# Ready-to-drink beverage (RTD) consumption in Thai population: trend and associated factors

**DOI:** 10.1101/2021.01.22.21250292

**Authors:** Udomsak Saengow, Ruttiya Asksonthong, Apinya Laohaprapanon

## Abstract

**Objectives:** To examine ready-to-drink beverage (RTD) consumption and to investigate the effects of gender and age on RTD consumption by using data 2011 and 2014 waves of a national alcohol survey.

**Design:** Analysis of data from Smoking and Drinking Behavior Survey (SADBeS) 2011 and 2014, a nationally representative survey.

**Setting:** Thailand

**Participants:** 177,350 (2011 survey) and 25,758 (2014 survey) samples of Thais aged 15 years or older who were randomly selected using multistage-sampling technique.

**Primary outcome:** RTD consumption in past 12 months (yes/no) as stated by survey participants

**Results:** The prevalence of RTD drinkers increased from 0.5% (95% CI, 0.5-0.5%) in 2011 to 2.4% (95% CI, 2.1-2.6%) in 2014. Female drinkers were 5.1 (95% CI, 4.1-6.4) times more likely to consume RTDs than male drinkers. The likelihood of drinking RTDs decreased with age. Drinking initiation before the legal purchasing age (20 years old) was associated with 1.5 (95% CI, 1.1-1.9) times likelihood of RTDs consumption.

**Conclusions:** A substantial increase in RTD consumption was observed in Thailand, a middle-income country, during 2011-2014. The consumption was notable in youths and females. Given that RTDs have been introduced into the Thai market relatively recently, this may be a part of the alcohol industry strategies to boost their sales in middle-income countries. Growth in RTD consumption could pose a challenge for health authorities to control alcohol-related harms in the future especially among youths and females.

**Article Summary:** *Strengths and limitations of this study:*

- A reliable estimate of prevalence of RTD consumption was obtained by using data from two waves of a large national representative survey.
- The lack of information about pattern of RTD consumption including quantity and drinking frequency limited further analysis.

*Funding:* This work was supported by the Center for Alcohol Studies, Thailand, grant number 62-02029-0043.

*Competing interests statement:* None declared.

## Introduction

Ready-to-drink beverages (RTDs) are pre-mixed beverages consisting of spirits, beer, or wine and a non-alcoholic base. This type of alcoholic beverage, which is also known as alcopops, is usually sweetened to obtain a pleasant taste ^1,2^. RTDs typically contain 5% to 9% of alcohol ^3^. Recently, RTDs have gained popularity in some countries and decreased in popularity in others. During 2006-2010, spending on RTDs in EU increased in the UK and Denmark, decreased in Ireland and Finland, and was constant in other EU countries ^4^. In Australia, comparing the 2017-2018 period with the 2016-2017 period, there was a 7.0% increase in total pure alcohol from RTD consumption; the increase was 5.5% in terms of per capita consumption ^5^.

The target consumers of RTDs are youths and females ^1,2^. Prevalence of youth and female RTD drinkers increased in high-income countries such as Australia, New Zealand, Switzerland, and the United States during 2007-2008 ^6-9^. A study from New Zealand found that RTDs were most popular among females and youths aged 14-17 years old ^6^. A study among Australian youths showed that RTDs were more appealing to younger age groups (under 18 years old). Moreover, RTDs were the most popular alcoholic beverages for first-time drinkers in a 12-13 years age group ^8^. Among Finnish teenagers, girls had a 33% higher chance of consuming RTDs in the latest drinking occasion compared to boys ^10^.

The packaging of RTDs evidently appeals to youths. A study assessing the effect of beverage product packaging on palatability rating reported that the packaging of RTDs appealed to younger age groups with a higher magnitude of effect observed in females. In contrast, the packaging of other alcoholic beverages appealed to older age groups ^11^. The price of RTDs is generally cheaper than other alcoholic beverages ^7^. Thus, the packaging and price of RTDs might attract youths to start drinking.

Existing evidence on RTD consumption is mostly from high-income and western countries. However, evidence about RTD consumption from middle-income countries is lacking. As patterns of alcohol consumption differ between countries with different income levels and cultures ^12,13^, data from low- and middle-income countries could provide a wider insight into RTD drinking behavior. This study aimed to examine RTD consumption and to investigate the effects of gender and age on RTD consumption by using data from two waves of a national alcohol survey in Thailand.

## METHODS

### Study design

This study analyzed data from two waves (2011 and 2014) of the national alcohol survey in Thailand to investigate RTD drinking behavior among the Thai population. The study protocol was approved by the Human Research Ethics Committee of Walailak university (WU-EC-MD-4-014-62).

### Data source

The data source of this study was the 2011 and 2014 waves of the Smoking and Drinking Behavior Survey (SADBeS). The SADBeS was conducted by the National Statistical Office, Thailand. The sampling scheme of the SADBeS was a stratified two-stage sampling. In the first stage, enumeration areas—which were used by the National Statistical Office for national census—were randomly selected with probability proportional to size from five strata: Bangkok, Central region (excluding Bangkok), Northern region, Northeastern region, and Southern region. In the second stage, households in the selected enumeration areas were chosen using a stratified sampling method. In the 2011 survey, all eligible persons in selected households were invited to be survey participants in this study. In the 2014 survey, one person per household was randomly selected from eligible household members to be a participant in this study. The total number of participants was 177,350 (from 66,300 households) from the 2011 survey and 25,758 (from 25,758 households) from the 2014 survey. The inclusion criteria included 15 years of age or older and fluent in Thai. Those with citizenships other than Thai were excluded. After being informed about the survey protocol, verbal consent was obtained from survey participants. For those aged 15-17, consent had to be provided by both participants and their parent (or guardian).

### Questionnaire

The questionnaire used in the survey consisted of several variables related to drinking behavior. The variables selected for this analysis included RTD consumption, gender, age, level of educational attainment, monthly income, drinking status, age of drinking initiation, and most frequented drinking venues. Data was cleaned and each variable was recoded to use in this analysis as follows.

The primary outcome variable was RTD consumption in the past 12 months (yes or no). The legal alcohol purchasing age in Thailand is 20 years or older ^14^. Hence, a dichotomous variable—drinking initiation before legal purchasing age—was generated by categorizing age of drinking initiation using a cut-off point of 20 years. The age of participants was grouped into four categories (i.e., 15-19, 20-24, 25-59, and 60+ years)

The level of educational attainment was classified into elementary level or lower, secondary level, and bachelor’s degree or beyond. Monthly income was classified into five quintiles: Quintile 1 was the least affluent group and Quintile 5 was the richest group. For the drinking status, those who drank within the past 12 months before the interview were considered current drinkers; regular drinkers were current drinkers who drank at least once a week; and occasional drinkers were current drinkers with less frequent drinking. The most frequented drinking venues were categorized as own home/friend’s home, restaurant/pub/bar, and other venues.

### Statistical analysis

All analysis in this study was weighted to represent the Thai general population aged 15 years or above. Data analysis was performed by R Version 3.5.1 ^15^ and ‘survey’ package was used in all weighted analysis ^16^. Graphs were generated using GraphPad Prism Version 4.0 for Windows (GraphPad Software, La Jolla California, USA, www.graphpad.com). Percentages and means were computed to describe survey participants. Prevalence of RTD drinkers stratified by each characteristic was estimated among the general population and among current drinkers. To determine factors associated with RTD consumption, multivariate logistic regression was used. The multivariate analysis was conducted to determine the trend in RTD consumption (year of survey), demographic characteristics of RTD drinkers (gender and age), socioeconomic status of RTD drinkers (education and income), and drinking-related factors (drinking status, age of drinking initiation, and drinking venues). Since some of these factors were only relevant for drinkers, data used in the regression analysis was limited to current drinkers. In this study, a p-value less than 0.05 was considered statistically significant.

## RESULTS

### Characteristics of the survey population

The estimated percentage of females in the survey population was 51.4% and 51.6% in 2011 and 2014, respectively. Most of the survey population was in the 25-59 years age group (more than 60% in both waves). About a half had their highest level of education at the elementary level or lower. Prevalence of current drinkers was 31.5% in 2011 and 32.3% in 2014, where an increase in the prevalence was from occasional drinkers. Statistics about drinking initiation was somewhat similar in both years. More than two-thirds in both years drank at home most frequently as shown in Table 1.

**Table 1.**
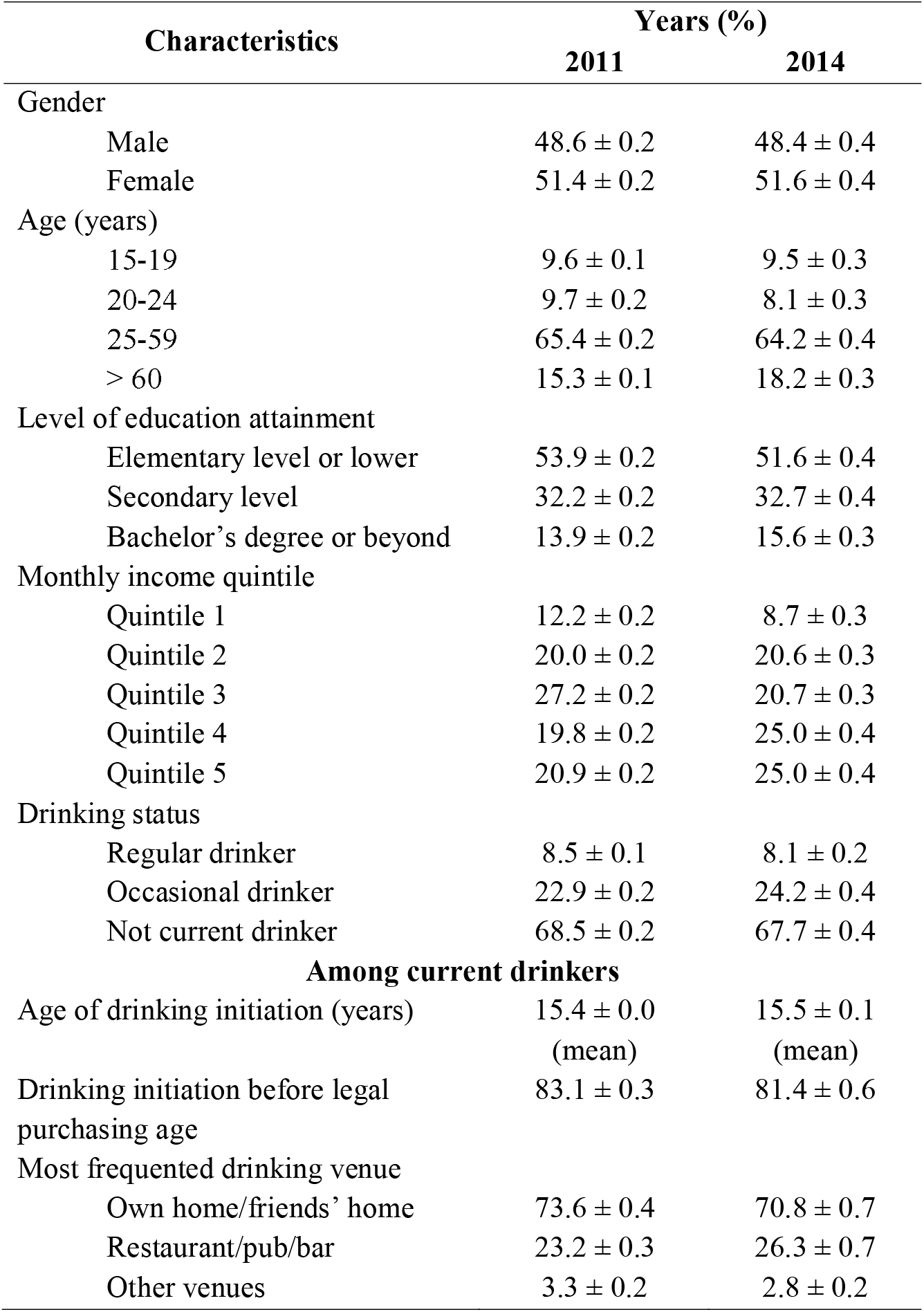
Characteristics of the survey population in 2011 and 2014 (weighted)

### Prevalence of RTD consumption

The estimated numbers of current drinkers were 16,992,017 (95% confidence interval [CI] 16,728,496-17,255,538) and 17,705,123 (95% CI 17,193,903-18,216,345) in 2011 and 2014, respectively. Among current drinkers, 270,606 (95% CI 233,083-308,130) in 2011 and 1,292,890 (95% CI 1,143,937-1,441,843) in 2014 drank RTDs. In both waves, the prevalence of RTD consumption was higher among females in 2011 and higher among males in 2014. The prevalence of RTD consumption in both genders increased considerably during the study period (Figure 1).

**Fig 1.**
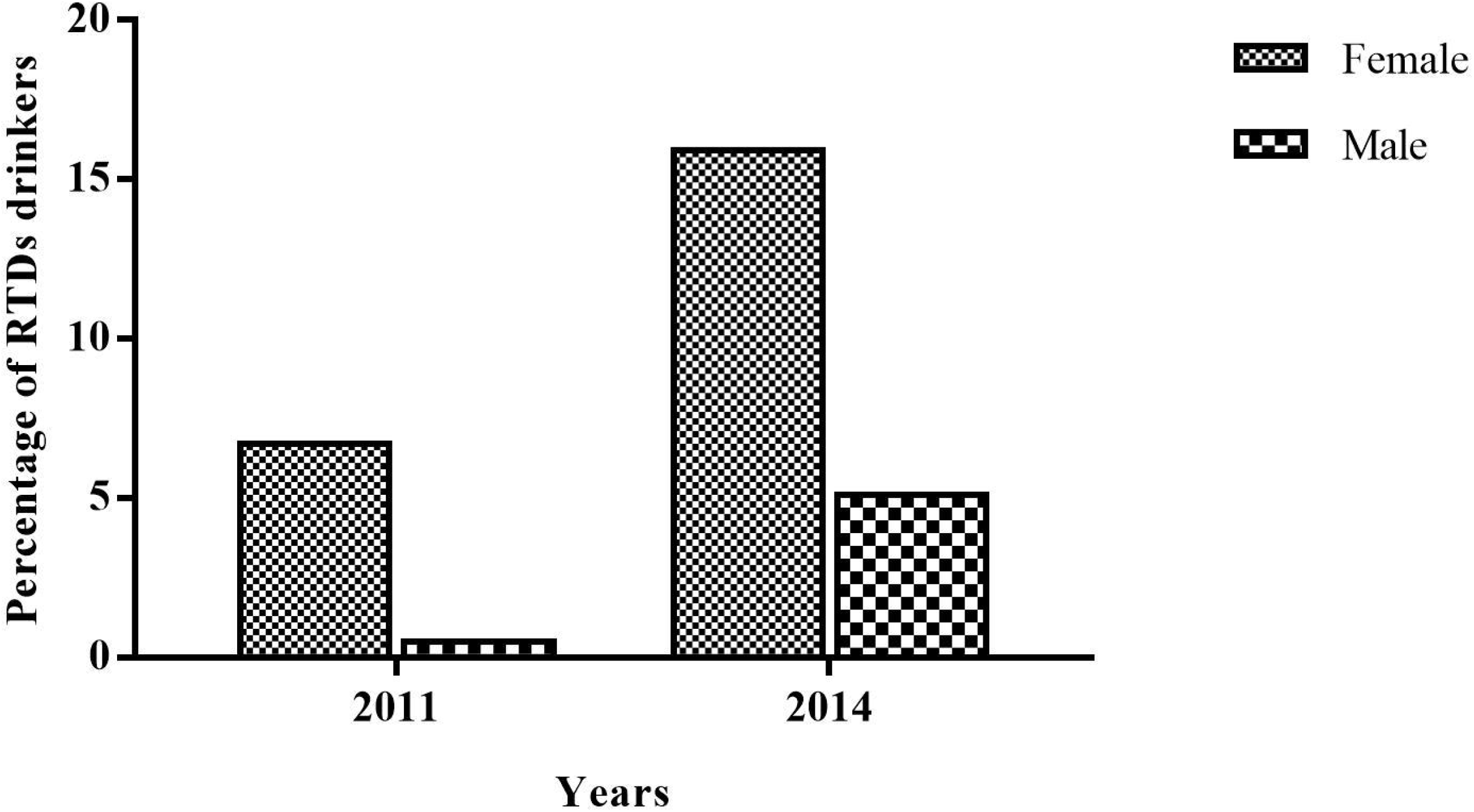

The age group with the highest proportion of RTD drinkers was 20-24 years old. The prevalence was lowest in the oldest age group. An increase in prevalence was observed in all age groups (Figure 2). Income and education level seemed to have a positive association with RTD consumption. Occasional drinkers had a higher prevalence of RTD consumption than regular drinkers. Those who most frequently drank at restaurants, bars, or pubs had the highest proportion of RTD consumption (Table 2).

**Fig 2.**
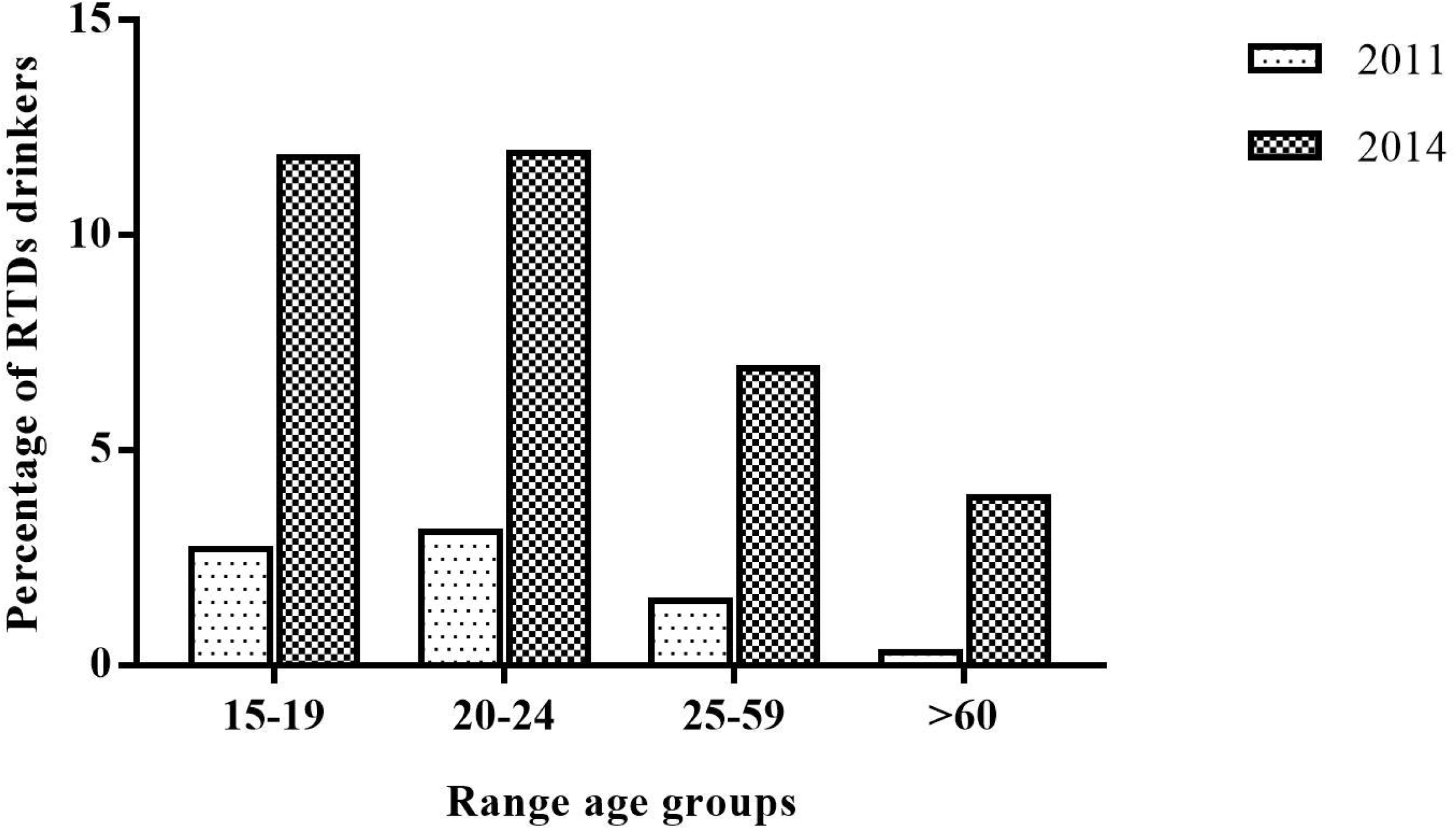

**Table 2.** Prevalence of RTD consumption in the general population and among current drinkers stratified by characteristics in 2011 and 2014 (weighted)

| Characteristics | General population (%) |  | Current drinkers (%) |  |
| --- | --- | --- | --- | --- |
|  | 2011 | 2014 | 2011 | 2014 |
| Total | 0.5 ± 0.0 | 2.4 ± 0.1 | 1.6 ± 0.1 | 7.3 ± 0.4 |
| Gender |  |  |  |  |
| Male | 0.3 ± 0.4 | 2.7 ± 0.2 | 0.5 ± 0.1 | 5.1 ± 0.4 |
| Female | 0.7 ± 0.6 | 2.1 ± 0.2 | 6.7 ± 0.5 | 15.9 ± 1.2 |
| Age (years) |  |  |  |  |
| 15-19 | 0.4 ± 0.1 | 2.1 ± 0.4 | 2.7 ± 0.7 | 11.8 ± 2.3 |
| 20-24 | 1.0 ± 0.2 | 4.0 ± 0.7 | 3.1 ± 0.6 | 11.9 ± 2.0 |
| 25-59 | 0.6 ± 0.4 | 2.7 ± 0.2 | 1.5 ± 0.1 | 6.9 ± 0.5 |
| > 60 | 0.0 ± 0.0 | 0.7 ± 0.2 | 0.3 ± 0.1 | 3.9 ± 0.9 |
| Level of education attainment |  |  |  |  |
| Elementary level or lower | 0.2 ± 0.0 | 1.6 ± 0.2 | 0.8 ± 0.1 | 5.1 ± 0.5 |
| Secondary level | 0.7 ± 0.1 | 2.9 ± 0.3 | 2.1 ± 0.2 | 8.4 ± 0.8 |
| Bachelor degree or beyond | 1.1 ± 0.1 | 3.7 ± 0.4 | 3.4 ± 0.4 | 12.0 ± 1.3 |
| Monthly income quintile |  |  |  |  |
| Quintile 1 | 0.1 ± 0.0 | 0.8 ± 0.3 | 1.8 ± 0.7 | 5.1 ± 1.7 |
| Quintile 2 | 0.3 ± 0.1 | 1.0 ± 0.2 | 1.3 ± 0.4 | 5.3 ± 1.0 |
| Quintile 3 | 0.4 ± 0.1 | 2.8 ± 0.3 | 1.1 ± 0.2 | 8.0 ± 0.9 |
| Quintile 4 | 0.7 ± 0.1 | 2.6 ± 0.3 | 1.7 ± 0.2 | 6.5 ± 0.7 |
| Quintile 5 | 0.9 ± 0.1 | 3.4 ± 0.4 | 2.3 ± 0.2 | 8.8 ± 0.9 |
| Drinking status |  |  |  |  |
| Regular drinker | NA | NA | 0.8 ± 0.2 | 2.9 ± 0.5 |
| Occasional drinker | NA | NA | 1.9 ± 0.1 | 8.8 ± 0.5 |
| Drinking initiation before legal purchasing age |  |  |  |  |
| Yes | NA | NA | 1.5 ± 0.1 | 7.3 ± 0.5 |
| No | NA | NA | $2.1 \pm 0.3$ | $7.2 \pm 0.8$ |
| Most frequented drinking venue |  |  |  |  |
| Own home/friends' home | NA | NA | $1.1 \pm 0.1$ | $6.2 \pm 0.5$ |
| Restaurant/pub/bar | NA | NA | $3.2 \pm 0.3$ | $10.3 \pm 0.9$ |
| Other venues | NA | NA | $1.9 \pm 0.6$ | $6.7 \pm 2.7$ |

### Factors associated with RTD consumption among current drinkers

Multiple logistic regression analysis was employed to examine determinants of RTD consumption among current drinkers. Drinkers were 4.6 times more likely to drink RTDs in 2014 than in 2011. A likelihood of drinking RTDs for females was five times higher than for males. Younger age groups had a higher chance of drinking RTDs. RTD consumption was more prevalent among the high education or high-income group. RTDs were more popular among occasional drinkers than among regular drinkers. Those who started drinking before the legal purchasing age were 48% more likely to drink RTDs (Table 3).

**Table 3.** Factors associated with RTD consumption among current drinkers.

| **Variables** | **Adj. OR (95% CI)** | **P-value** |
| --- | --- | --- |
| Year (ref: 2011) |  |  |
| 2014 | 4.6 (3.8, 5.6) | < 0.001* |
| Gender (ref: Male) |  |  |
| Female | 5.1 (4.1, 6.4) | < 0.001* |
| Age (ref: > 60) |  |  |
| 15-19 | 3.0 (1.6, 5.7) | 0.001* |
| 20-24 | 2.2 (1.2, 3.9) | 0.011* |
| 25-59 | 1.3 (0.8, 2.0) | 0.365 |
| Level of education attainment (ref: Elementary level or lower) |  |  |
| Secondary level | 1.4 (1.1, 1.8) | 0.019* |
| Bachelor's degree or beyond | 2.1 (1.6, 2.9) | < 0.001* |
| Monthly income quintile (ref: Quintile 1) |  |  |
| Quintile 2 | 1.4 (0.7, 2.7) | 0.353 |
| Quintile 3 | 2.2 (1.2, 4.2) | 0.015* |
| Quintile 4 | 2.1 (1.1, 3.9) | 0.026* |
| Quintile 5 | 2.6 (1.4, 5.0) | 0.003* |
| Drinking status (ref: Regular drinker) |  |  |
| Occasional drinker | 1.9 (1.4, 2.7) | < 0.001* |
| Age of drinking initiation before the legal purchasing age (ref: No) |  |  |
| Yes | 1.5 (1.1, 1.9) | 0.004* |
| Most frequented drinking venue (ref: Own home/friends' home) |  |  |
| Restaurant/pub/bar | 1.1 (0.9, 1.4) | 0.329 |
| Other venues | 1.3 (0.7, 2.6) | 0.405 |

\* $p$ -value < 0.05.

## DISCUSSION

RTD drinking behavior among the Thai population aged 15 years and above was investigated using the data from the SADBeS (2011 and 2014). We found that the trend in RTD consumption between 2011 and 2014 was increasing. Females drank RTDs more than males. RTDs were popular among younger age groups, persons with high education, persons with high income, and occasional drinkers. Early drinking initiation was also associated with RTD consumption.

The finding that Thai females had a significantly higher chance of being RTD drinkers than Thai males was similar to the evidence from high-income western countries. A study among New Zealand youths and young adults revealed that females aged 14-17 years old was a group with the highest prevalence of RTD consumption ^6^. Similarly, Swedish young females had a higher proportion of RTD consumption than young males ^7^. In Australia, RTDs were the most popular alcoholic beverage for young females ^17^.

Younger age was an important determinant of RTD consumption. Our findings suggested that persons aged < 25 years old had a significantly higher chance of drinking RTDs as compared to those aged >60 years old and drinking initiation before the legal purchasing age was associated with drinking RTDs. A survey from New Zealand showed that the age group with the highest prevalence of RTDs drinkers was 16-19 years old, and the prevalence linearly decreased with age ^6^. A study among Australian youths found that RTDs were the most popular alcoholic beverages among respondents aged 12-17 years old ^17^. A survey conducted a few months after RTDs were first introduced into the Swedish alcohol market found that 50% of the age group 15-18 years old had already tried RTDs. In a survey conducted a year later, that figure increased to 70% ^18^.

In the 1990s, RTDs were introduced to the market as a new type of alcoholic beverage ^19^. It has been primarily designed to attract young drinkers. To achieve that, some RTDs have fruit flavors and sweet taste added; others have the taste of alcohol removed from their drinks. The price of RTDs is intentionally set to be cheaper than other alcoholic drinks ^20^. An early study on RTD perception by youths demonstrated that RTDs were particularly attractive to youths aged <16 years old ^21^. The design of RTD packaging boosted its palatability rating among youths and females ^11^. Correspondingly, early drinking initiation was associated with the consumption of RTDs in our study. RTDs accounted for an increase in alcohol consumption among young boys and girls ^22^.

Our findings suggested that there was an increasing trend of RTD consumption in Thailand during the study period. Unlike their counterparts in western countries, less than 20% of Thai teenagers (aged 15-19 years old) have been current drinkers. The proportion of current drinkers among Thai teenagers was 14.0% in 2011 and 14.6% in 2014 (calculated from the SADBeS 2011 and 2014 data). Therefore, the market for alcoholic beverages in Thailand is far from saturated. This provides an opportunity for the alcohol industry to boost their future sales. Furthermore, a multinational alcohol company has expected growth in the demand for alcoholic beverages in the Asia-Pacific region from an increasing number of middle-class consumers ^23^. Data from a Thai major alcoholic beverage company shows that 5 out of their 10 RTD products have been introduced to the market after 2011 (during and after our study period), and 2 were introduced in 2009 (2 years before our study period) ^24^. Marketing and promotion activities linked to the introduction of new RTD products may cause an increase in RTD consumption in Thailand. Imposing taxes on RTDs could help cut its consumption ^25^.

Consumption of RTDs was more prevalent in higher income and higher education groups. Our findings were in agreement with the results of the study of European countries ^4^. An association between RTD consumption and drinking frequency tended to be negative. A study from New Zealand found a negative relationship between RTD consumption and drinking frequency among an overall sample ^6^. The relationship was not statistically significant in an analysis of Finnish surveys ^10^. In our study, occasional drinkers were more likely to drink RTDs.

A major strength of this study was that we used data from a large population-based survey. With weighted analysis, we were able to obtain a reliable estimate of the prevalence of RTD consumption. The lack of detailed data about RTD drinking behavior, such as drinking frequency and drinking quantity, limited further analysis. Growth in RTD consumption could pose a challenge for health authorities with respect to an increase in alcohol consumption among youths and females. Further studies to obtain insight about the time, place, and circumstance of RTD consumption are worth considering.

## Data Availability

Authors obtained the data used in this study from the Center for Alcohol Studies with the permission under the contract of this study. Request for the data can be made to the corresponding author. Access to the data must be permitted by the Center for Alcohol Studies.

## Author contributions

US obtained the funding and designed the study. US, RA and AL analyzed the data and interpreted the results. RA and AL were responsible for the first draft of manuscript. US worked on subsequent and final drafts. All authors critically reviewed and approved the manuscript.

